# Characterizing the polygenic architecture of complex traits in populations of East Asian and European descent

**DOI:** 10.1101/2023.05.25.23290542

**Authors:** Antonella De Lillo, Frank R. Wendt, Gita A. Pathak, Renato Polimanti

## Abstract

To investigate the polygenicity of complex traits in populations of East Asian (EAS) and European (EUR) descents, we leveraged genome-wide data from Biobank Japan, UK Biobank, and FinnGen cohorts. Specifically, we analyzed up to 215 outcomes related to 18 health domains, assessing their polygenic architecture via descriptive statistics, such as the proportion of susceptibility SNPs per trait (π_c_). While we did not observe EAS-EUR differences in the overall distribution of polygenicity parameters across the phenotypes investigated, there were ancestry-specific patterns in the polygenicity differences between health domains. In EAS, pairwise comparisons across health domains showed enrichment for π_c_ differences related to hematological and metabolic traits (hematological fold-enrichment=4.45, p=2.15×10^−7^; metabolic fold-enrichment=4.05, p=4.01×10^−6^). For both categories, the proportion of susceptibility SNPs was lower than that observed for several other health domains (EAS-hematological median π_c_=0.15%, EAS-metabolic median π_c_=0.18%) with the strongest π_c_ difference with respect to respiratory traits (EAS-respiratory median π_c_=0.50%; Hematological-p=2.26×10^−3^; Metabolic-p=3.48×10^−3^). In EUR, pairwise comparisons showed multiple π_c_ differences related to the endocrine category (fold-enrichment=5.83, p=4.76×10^−6^), where these traits showed a low proportion of susceptibility SNPs (EUR-endocrine median π_c_=0.01%) with the strongest difference with respect to psychiatric phenotypes (EUR-psychiatric median π_c_=0.50%; p=1.19×10^−4^). Simulating sample sizes of 1,000,000 and 5,000,000 individuals, we also showed that ancestry-specific polygenicity patterns translate into differences across health domains in the genetic variance explained by susceptibility SNPs projected to be genome-wide significant (e.g., EAS hematological-neoplasm p=2.18×10^−4^; EUR endocrine-gastrointestinal p=6.80×10^−4^). These findings highlight that traits related to the same health domains may present ancestry-specific variability in their polygenicity.

## INTRODUCTION

Genome-wide association studies (GWAS) are improving our understanding of the predisposition to human traits and diseases, providing insights into their underlying biological mechanisms (1). However, their ability to disentangle complex phenotypes is mainly proportional to the sample size of the cohorts investigated, because most human traits and diseases are characterized by a polygenic architecture (i.e., their heritability is due to the contribution of thousands of risk loci with small individual effects) (2-4). There are differences in the degree of polygenicity among complex traits where extremely high polygenicity is observed for psychiatric disorders such as depression and relatively low polygenicity is present for physical conditions such as ulcerative colitis (5). Multiple mechanisms are likely to contribute to this variation. Purifying selection (i.e., the selective removal of deleterious alleles across the genome) plays a major role in shaping the polygenic architecture of human traits and diseases (6, 7). However, phenotypic heterogeneity could also contribute to the degree of polygenicity observed. Among psychiatric disorders, the number of diagnostic combinations was associated with effect size variance for trait-associated loci (8). Understanding the dynamics shaping the polygenicity variation across the human phenotypic spectrum can generate important insights into the evolutionary basis of human traits. Additionally, defining polygenicity patterns can help to estimate more accurately the statistical power of phenotype-specific gene discovery analyses and to model more precisely polygenic scores (PGS) to stratify disease risk. Unfortunately, the majority of studies investigating polygenicity patterns across human traits and diseases are based on data generated from individuals of European descent (5-8). Although we expect a convergence in the biology of complex phenotypes among worldwide populations, there may be differences due to the evolutionary history of ancestry groups.

In the present study, we leveraged genome-wide association statistics available from Biobank Japan (BBJ) (9), UK Biobank (UKB) (10), and FinnGen (11) to estimate the effect size distribution of multiple traits comparing differences among health domains in individuals of East Asian and European descent (EAS N=178,726 and EUR N=492,803, respectively; Supplemental Table 1). In addition to estimating the number of susceptibility single nucleotide polymorphisms (SNPs) and their effect size distribution, we also calculated the genetic variance explained by the genome-wide significant variants projected considering sample sizes of 1,000,0000 and 5,000,000 individuals. Our findings provide novel information regarding polygenicity patterns across the human phenotypic spectrum, highlighting possible similarities and differences between EAS and EUR ancestries.

## RESULTS

### Effect-Size Distribution Analysis

We analyzed genome-wide association statistics previously generated from BBJ, and a UKB-FinnGen meta-analysis (12). These included up to 215 traits related to 18 health domains (Table 1; Supplemental Table 1). Using GENESIS (GENetic Effect-Size distribution Inference from Summary-level data) approach (5), we estimated the proportion of susceptibility SNPs per trait (π_c_), the variance parameter for non-null SNPs (σ^2^), and residual effects not captured by the variance of effect sizes (a) (Table 1; Supplemental Table 1). The analyses were conducted separately for EAS and EUR. To determine within- and between-population differences of π_c_, σ^2^, and a parameters, we applied the non-parametric Kruskal–Wallis (KW) test and conducted post-hoc analyses using Dunn non-parametric test for the pairwise comparisons.

**Table 1:** Distribution (median, maximum, and minimum) of polygenicity parameters across phenotypic categories in populations of East Asian and European descent (EAS and EUR, respectively). Estimates are reported as percentages. π_c_: the proportion of susceptibility SNPs per trait; σ^2^: the variance parameter for non-null SNPs; *a*: residual effects not captured by the variance of effect sizes.

| Category | EAS, Median (Max – Min) |  |  |  | EUR, Median (Max – Min) |  |  |  |
| --- | --- | --- | --- | --- | --- | --- | --- | --- |
| | N | %$\pi_c$ | %$\sigma^2$ | %$a$ | N | %$\pi_c$ | %$\sigma^2$ | %$a$ |
| Body Structures | 4 | 0.71 (1.01 – 0.47) | 0.002 (0.01 – 0.001) | 0.0002 (0.0003 – 0.0001) | 1 | 0.50 | <0.0001 | 0.0002 (0.0002) |
| Cardiovascular | 22 | 0.50 (0.67 – 0.06) | 0.0074 (0.08 – <0.0001) | 0.0001 (0.002 – -0.02) | 18 | 0.50 (0.50 – 0.17) | 0.01 (0.03 – <0.0001) | 0.0002 (0.001 – -0.002) |
| Dermatological | 4 | 0.50 (0.51 – 0.005) | <0.0001 (1.30 – <0.0001) | 0.0005 (11.07 – -0.01) | 4 | 0.50 (0.50 – 0.44) | 0.03 (0.04 – <0.0001) | -0.002 (-0.0002 – -0.02) |
| Ear, Nose, Throat | 5 | 0.50 (0.65 – 0.50) | 0.02 (0.06 – <0.0001) | -0.001 (0.001 – -0.01) | 5 | 0.50 (0.50 – 0.04) | 0.01 (0.21 – 0.01) | 0.0001 (0.001 – -0.001) |
| Endocrine | 5 | 0.50 (0.50 – 0.01) | 0.004 (0.89 – <0.0001) | 0.001 (0.002 – -0.0003) | 5 | 0.01 (0.14 – 0.005) | 0.81 (1.84 – 0.04) | 0.001 (0.002 – 0.0004) |
| Environment | 1 | 0.50 | 0.01 | -0.0002 | 1 | 0.50 | 0.005 | 0.0001 (0.0001) |
| Gastrointestinal | 8 | 0.50 (0.50 – 0.11) | 0.004 (0.06 – <0.0001) | 0.0001 (0.003 – -0.002) | 7 | 0.50 (1.07 – 0.10) | 0.003 (0.08 – <0.0001) | 0.0004 (0.001 – -0.0003) |
| Hematological | 18 | 0.15 (0.50 – 0.02) | 0.01 (0.03 – <0.0001) | 0.0001 (0.001 – -0.13) | 4 | 0.50 (0.50 – 0.50) | 0.004 (0.01 – <0.0001) | 0.0001 (0.0003 – -0.01) |
| Immunological | 28 | 0.50 (0.71 – 0.002) | 0.02 (1.01 – <0.0001) | <0.0001 (0.008 – -0.02) | 27 | 0.50 (0.67 – 0.00) | 0.01 (3.56 – <0.0001) | 0.0002 (0.002 – -0.06) |
| Medication | 22 | 0.50 (0.62 – 0.01) | 0.01 (0.32 – 0.001) | 0.0001 (0.001 – -0.003) | 0 | NA | NA | NA |
| Metabolic | 31 | 0.18 (0.52 – 0.002) | 0.01 (0.79 – <0.0001) | 0.0001 (0.001 – -0.004) | 15 | 0.50 (0.78 – 0.003) | 0.01 (4.26 – <0.0001) | 0.0004 (0.01 – -0.002) |
| Musculoskeletal | 9 | 0.50 (0.50 – 0.01) | <0.0001 (0.50 – <0.0001) | 0.0001 (0.005 – -0.04) | 9 | 0.50 (1.12 – 0.17) | 0.005 (0.04 – <0.0001) | 0.0002 (0.001 – -0.04) |
| Neoplasms | 17 | 0.50 (0.60 – 0.003) | 0.03 (3.19 – <0.0001) | 0.0002 (0.01 – -0.01) | 17 | 0.50 (0.50 – 0.002) | 0.02 (2.96 – <0.0001) | 0.0004 (0.003 – -0.003) |
| Neurological | 7 | 0.50 (0.52 – 0.32) | 0.01 (0.18 – <0.0001) | -0.001 (0.0004 – -0.02) | 6 | 0.50 (0.50 – 0.36) | 0.01 (0.02 – <0.0001) | -0.0004 (0.001 – -0.004) |
| Ophthalmological | 8 | 0.50 (0.50 – 0.08) | 0.005 (0.04 – <0.0001) | <0.0001 (0.002 – -0.002) | 7 | 0.20 (0.50 – 0.003) | 0.03 (1.98 – <0.0001) | 0.0005 (0.001 – -0.0002) |
| Psychiatric | 5 | 0.50 (0.67 – 0.50) | 0.02 (0.18 – <0.0001) | -0.002 (0.001 – -0.02) | 5 | 0.50 (0.90 – 0.50) | 0.01 (0.03 – 0.01) | 0.0002 (0.0003 – -0.001) |
| Respiratory | 9 | 0.50 (0.50 – 0.50) | <0.0001 (0.02 – <0.0001) | -0.001 (0.001 – -0.01) | 9 | 0.50 (0.84 – 0.02) | 0.01 (0.21 – 0.002) | -0.0003 (0.001 – -0.001) |
| Urogenital | 12 | 0.50 (0.55 – 0.03) | 0.0004 (0.15 – <0.0001) | -0.0002 (0.003 – -0.01) | 12 | 0.50 (0.50 – 0.01) | 0.01 (0.14 – <0.0001) | 0.0003 (0.000 – -0.0002) |

For the within-population analyses, we investigated whether there are differences among traits related to different phenotypic categories (Table 1; Supplemental Table 1). This analysis was limited to categories including at least four traits. In EAS, the proportion of susceptibility SNPs was statistically different among phenotypic categories (π_c_; KW statistic=39.14, p=6.41×10^−3^), while no difference was observed with respect to σ^2^ and a parameters (Supplemental Table 2). Although they did not survive false discovery rate correction (FDR) for multiple testing (Supplemental Table 3), the nominally significant pairwise comparisons showed enrichment for π_c_ differences related to hematological and metabolic traits (hematological fold-enrichment=4.45, p=2.15×10^−7^; metabolic fold-enrichment=4.05, p=4.01×10^−6^). For both categories, the proportion of susceptibility SNPs was lower than that observed for several other health domains (EAS-hematological median π_c_=0.15%, EAS-metabolic median π_c_=0.18%; Figure 1) with the strongest π_c_ difference with respect to respiratory traits (EAS-respiratory median π_c_=0.50%; Hematological Dunn statistic=3.05, p=2.26×10^−3^; Metabolic Dunn statistic=2.92, p=3.48×10^−3^).

**Figure 1:**
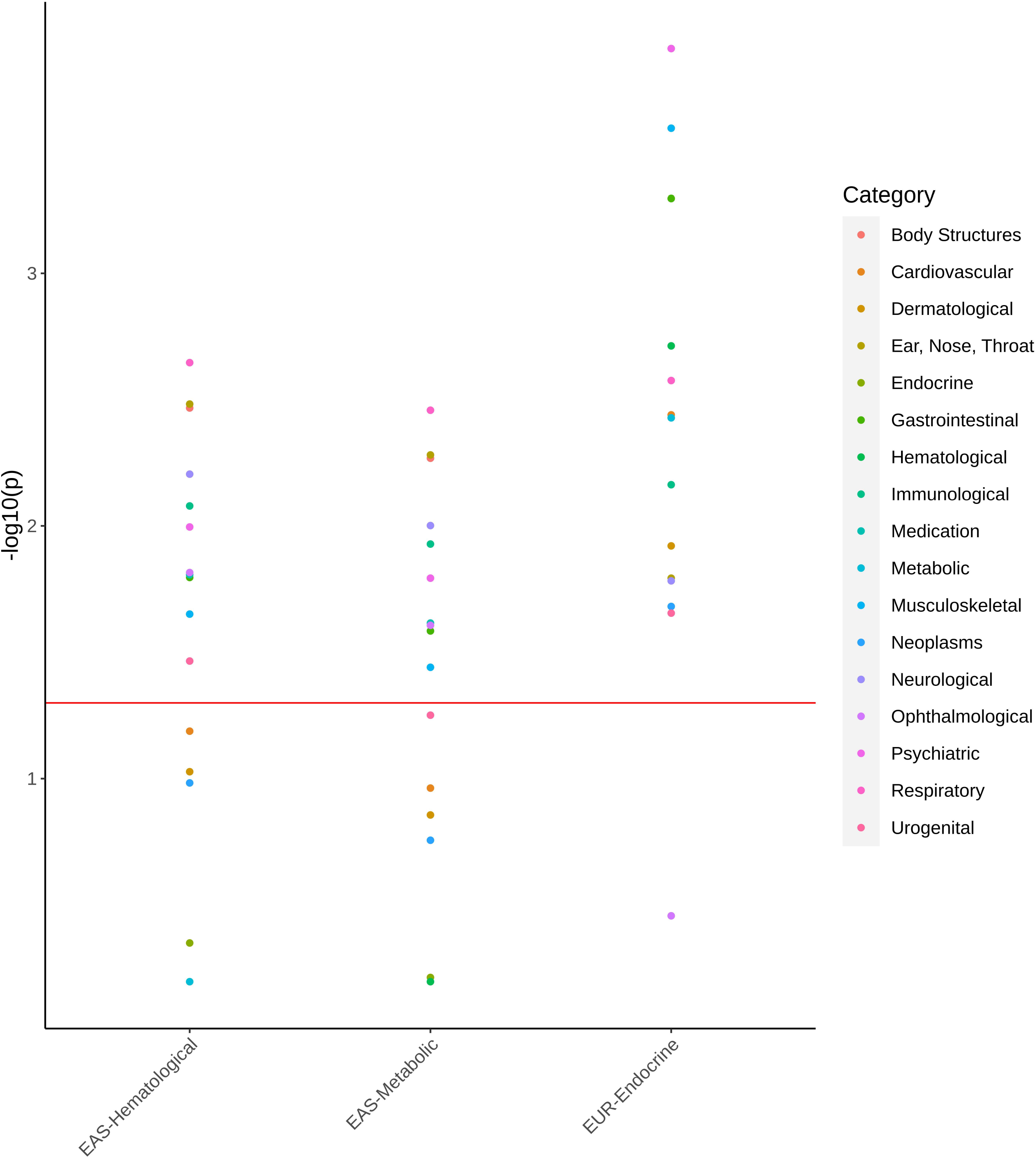
Pair-wise comparison (Dunn test) of the proportion of susceptibility SNPs (π_c_) in hematological and metabolic traits assessed in populations of East Asian descent (EAS-Hematological and EAS-metabolic, respectively) and in endocrine traits assessed in population of European descent (EUR-Endocrine) with respect to other phenotypic categories. Details of each comparison are available in Supplemental Tables 3 and 4.

In EUR, significant differences among phenotypic categories were observed with respect to π_c_ (KW statistic=35.27, p=0.013) and a parameters (KW statistic=31.17, p=0.039), but not for σ^2^ (KW statistic=25.05, p=0.477). For π_c_, nine pairwise comparisons showed differences surviving multiple testing correction (FDR q<0.05; Supplemental Table 4). Seven of them were related to the endocrine category (fold-enrichment=5.83, p=4.76×10^−6^), where these traits showed a low proportion of susceptibility SNPs (EUR-endocrine median π_c_=0.01%; Figure 1) with the strongest difference with respect to psychiatric phenotypes (EUR-psychiatric median π_c_=0.50%; Dunn statistic=3.83, p=1.19×10^−4^). For parameter a, we observed four pairwise comparisons surviving FDR correction (FDR q<0.05; Supplemental Table 5). Three of them were related to the endocrine category (EUR-endocrine median a=0.001%) that showed higher residual effects than dermatological traits (EUR-dermatological median a=-0.002%; Dunn statistic=-3.91, p=9.32×10^−5^), respiratory phenotypes (EUR-respiratory median a=-0.0003%; Dunn statistic=-3.38, p=7.17×10^−4^), and neurological outcomes (EUR-neurological median a=-0.0004%; Dunn statistic=-3.30, p=9.54×10^−4^). The dermatological category also showed lower residual effects than metabolic traits (Dunn statistic=3.20, p=1.39×10^−3^).

Comparing EAS and EUR, we observed a statistically significant difference with respect to residual effects not captured by the variance of effect sizes (parameter a: KW statistic=8.79, p=3.03×10^−3^), but not for π_c_ and σ^2^ (Supplemental Table 6). A category-specific EAS-EUR comparison of parameter a (Supplemental Table 7) highlighted nominally significant differences for metabolic traits (EAS median a=0.0001%; EUR median a=0.0004%; KW statistic=5.22, p=0.022) and urogenital phenotypes (EAS median a=-0.0002; EUR median a=0.0003%; KW statistic=4.08, p=0.043). While we did not observe the EAS-EUR difference in the overall distribution of the proportion of susceptibility SNPs, the within-population analysis showed significant π_c_ variation across phenotypic categories in both EAS and EUR, there was no overlap among the pairwise differences (Supplemental Tables 3 and 4). Indeed, EAS π_c_ differences were enriched for the hematologic domain (fold-enrichment=4.45, p=2.15×10^−7^) and metabolic categories (fold-enrichment=4.05, p=4.01×10^−6^), while EUR π_c_ differences were enriched for endocrine category (fold-enrichment=5.83, p=4.76×10^−6^). There was no enrichment convergence also when considering nominally significant π_c_ differences in both populations. Testing EAS-EUR π_c_ differences within these three categories, hematologic traits in EAS showed a lower proportion of susceptibility SNPs than that observed in EUR (EAS median π_c_=0.15%, EUR median π_c_=0.50%, KW statistic=5.96, p=0.015. A similar trend was also observed for metabolic traits (EAS median π_c_=0.18%, EUR median π_c_=0.50%, KW statistic=3.33, p=0.068). Conversely, the endocrine category had a higher proportion of susceptibility SNPs in EAS (endocrine median π_c_=0.50%) compared to EUR (endocrine median π_c_=0.01%; KW statistic=4.03, p=0.045).

### Projected Genetic Variance Explained by Susceptibility SNPs

To characterize further the implications of EAS and EUR polygenicity variation across phenotypic categories, we estimated the proportion of genetic variance explained by susceptibility SNPs reaching genome-wide significance considering projected sample sizes of 1,000,000 and 5,000,000 individuals (GV_%1M_ and GV_%5M_, respectively; Supplemental Table 8). For some traits, GV_%_ could not be calculated because of the low SNP-based heritability z-scores.

In EAS, both GV_%1M_ and GV_%5M_ showed significant differences across phenotypic categories (GV_%1M_ KW statistic=21.95, p=1.24×10^−3^; GV_%5M_ KW statistic=20.91, p=1.91×10^−3^). Results of the pairwise comparisons were consistent between the two analyses (i.e., N=1,000,000 and N=5,000,000) with five FDR-significant differences shared between them (FDR q<0.05; Figure 2, Supplemental Table 9) with the strongest one being between hematological and neoplasm categories (GV_%1M_ Dunn statistic=3.70, p=2.18×10^−4^; GV_%5M_ Dunn statistic=3.50, p=4.70×10^−4^).

**Figure 2:**
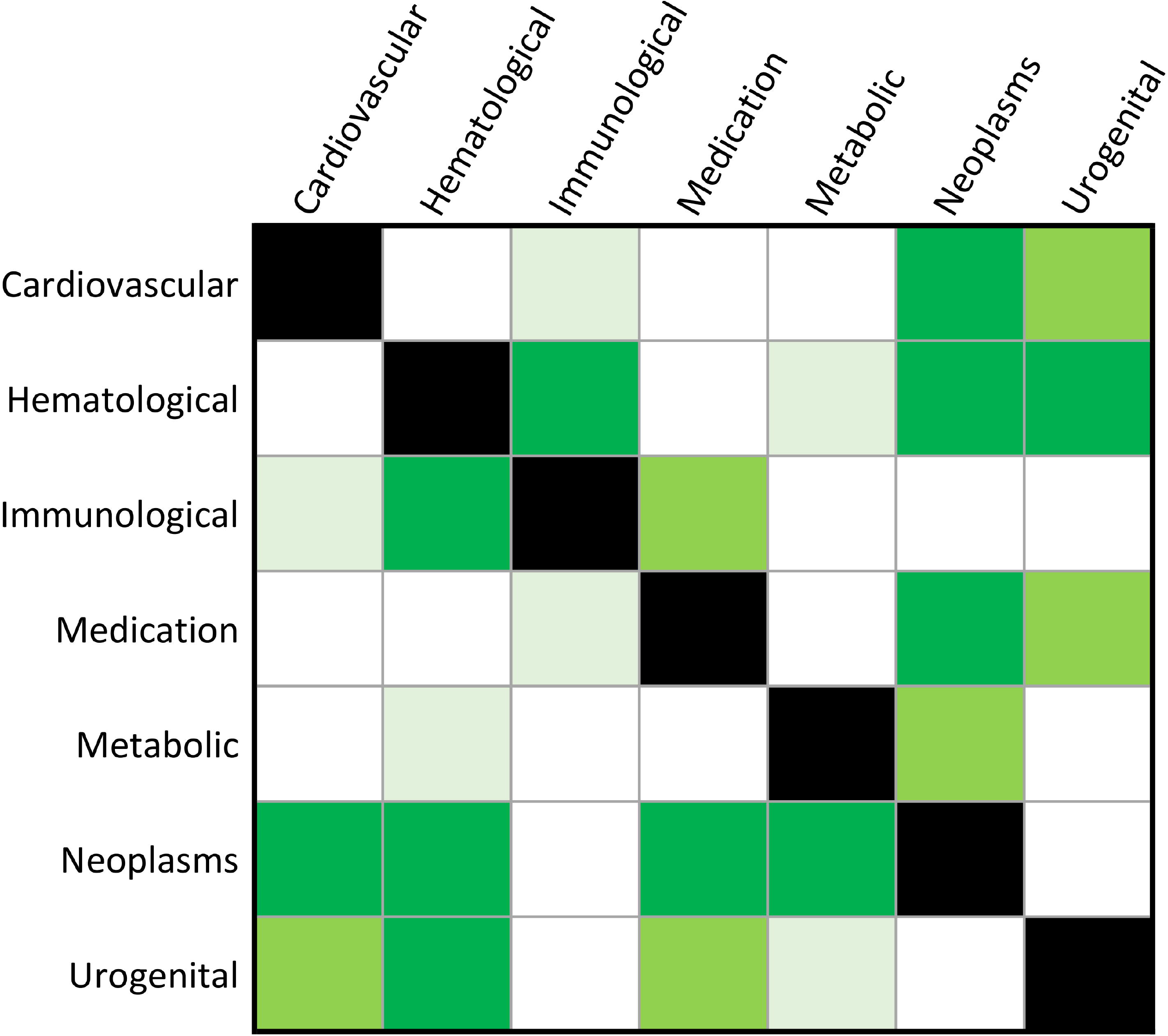
Pairwise comparison (Dunn test) among phenotypic categories of the proportion of genetic variance explained by susceptibility SNPs reaching genome-wide significance considering projected sample sizes of 1,000,000 (bottom triangle) and 5,000,000 individuals (upper triangle) of East Asian descent. Cell color corresponds to the statistical significance of the pair-wide comparison: bright green (false discovery rate q<0.05), green (p<0.05), light green (p<0.1), and white (p>0.1). Details of each comparison are available in Supplemental Table 9.

In EUR, while KW test showed differences across phenotypic categories with respect to two analyses (GV_%1M_ KW statistic=21.95, p=1.24×10^−3^; GV_%5M_ KW statistic=20.91, p=1.91×10^−3^), the only FDR-significant pair-wise difference was between endocrine and gastrointestinal categories (GV_%1M_ and GV_%5M_: Dunn statistic=-3.39, p=6.80×10^−4^; Supplemental Table 10)

In the between-population comparison, we did not find a significant EAS-EUR difference with respect to GV_%1M_ and GV_%5M_ estimates considering the overall distribution of the traits available and the major phenotypic categories (p>0.2; Supplementary Tables 11 and 12, respectively).

## DISCUSSION

The present study leveraged BBJ, UKB, and FinnGen cohorts to investigate the polygenicity of complex traits in EAS and EUR individuals. To our knowledge, this is the first effort to comprehensively investigate polygenicity patterns across the human phenotypic spectrum in multiple ancestry groups. In line with the expectation that the biology of complex phenotypes should converge among worldwide populations, we found that there was no statistical difference in the proportion of susceptibility SNPs when testing the overall distribution across multiple traits. However, when investigating polygenicity variation (i.e., the proportion of susceptibility SNPs per trait) with respect to specific health domains, we observed ancestry-specific patterns.

In EAS, there was strong enrichment for metabolic and hematologic categories when testing differences with respect to the proportion of susceptibility SNPs. Specifically, metabolic and hematologic traits showed a lower degree of polygenicity (i.e., a lower proportion of susceptibility SNPs) than phenotypes related to several other categories (e.g., respiratory, neurological, psychiatric, and immunological). Since the polygenicity of complex traits is primarily driven by purifying selection (6, 7), specific evolutionary dynamics may be responsible for the low polygenicity of metabolic and hematologic traits. Compared to other ancestry groups, metabolic risk in EAS populations appears to be stronger where individuals tend to develop prediabetes and diabetes at a younger age and at a lower body mass index and waist circumference (13, 14). The genetic risk of type-2 diabetes is partially shared between EAS and EUR, but there may be EAS-specific pathways related to skeletal muscle, adipose, and liver development and function (15). This ancestry-specific genetic risk may be due to adaptations to cereal-based diets. Indeed, the diet of EAS populations relied on wild and domesticated rice for more than 10,000 years (16, 17). This long exposure to a cereal associated with high glucose load may be responsible for the signatures of selective sweeps and polygenic adaption observed in Chinese, Korean, and Japanese populations in genes involved in fatty acids metabolism, cholesterol/triglycerides biosynthesis from carbohydrates, regulation of glucose homeostasis, and production of retinoic acid (18).

With respect to the lower polygenicity of hematologic traits in EAS compared to other health domains, there is limited information regarding which dynamics may be responsible. However, there is consistent literature regarding the impact of human evolutionary history on shaping the variation of genes related to hematologic phenotypes. For example, adaptation to malaria strongly influenced the genetics of hematologic traits through a systematic positive selection of protective alleles that is likely to be partially different across ancestry groups (19). Denisovan-introgressed alleles were responsible for high-altitude adaption in Tibetans, which showed a modified response to hypoxia-altering changes in hemoglobin concentration (20). Denisovan-introgressed alleles were also associated with hematologic traits (e.g., albumin-globulin ratio) in EAS (21). Blood biomarkers showed strong cross-ancestry heterogeneity in the effect of genome-wide significant loci (22). While these mechanisms do not directly explain the lower polygenicity of hematologic traits in EAS, they support that blood-related phenotypes played an important role in human evolution and that there may be specific adaptation processes in EAS populations affecting them.

In EUR, endocrine traits showed a lower polygenicity degree than other complex phenotypes (i.e., psychiatric, musculoskeletal, gastrointestinal, hematological, respiratory, cardiovascular, and metabolic traits). The endocrine system plays a key role in many aspects (e.g., development, reproduction, and response to the environment) that were essential to the success of the human species (23). Indeed, several studies reported evidence of signatures of multiple selective pressures acting in genes related to the endocrine system (24-26). In EUR, recent positive directional selection was observed in human male reproductive genes in response to different environmental conditions (27). A Neanderthal introgressed allele increasing the levels of progesterone receptor is associated with having more siblings, fewer miscarriages, and less bleeding during early pregnancy in EUR individuals (28). Dietary changes may have also contributed to shaping hormonal responses such as the effect of agriculture introduction on thyroid hormones (29). The presence of multiple adaption processes may have reduced the effect of purifying selection acting on endocrine-related genes in EUR.

Beyond the link between human evolution and the genetic of complex traits, the ancestry-specific polygenicity patterns have implications on the translation of genetic information into clinical care. Indeed, there are ongoing efforts into defining PGS to stratify disease risk (30). For certain health outcomes, PGS power to stratify individual risk appears to be comparable to some monogenic mutations (31). One of the main limitations in PGS application is modeling polygenic risk across ancestry groups due to the limited representativeness of worldwide populations among large-scale GWAS (32). While methods are being developed to perform effectively cross-population polygenic prediction across ancestry groups (33, 34), our findings highlight that modeling polygenicity may need to account for ancestry-specific differences across different health domains. Specifically, the genetic variance explained by susceptibility SNPs projected with respect to the sample sizes of 1,000,000 and 5,000,000 individuals shows how future GWAS may generate extremely powerful PGS with ancestry-specific predictivity variation with respect to certain phenotypic categories. Our results point to several such health domains that may require closer attention to cross-ancestry effects.

Most of our findings were related to the proportion of susceptibility SNPs per trait, which is informative of the polygenicity degree of the phenotypes investigated. However, we also identified significant between- and within-population differences in the residual effects not captured by the variance of effect sizes (i.e., parameter a). This reflects the potential systematic bias in variance estimates due to effects such as population stratification or cryptic relatedness (5). In line with the fact that current methods can adequately control these GWAS confounders, we observed extremely low estimates of parameter a (Table 1). Nevertheless, we saw differences between ancestries and between categories. This is likely due to the characteristics of the cohorts and/or the populations investigated. The between-category differences may be related to the specific dynamics linking population structure to the assessment of traits analyzed. As mentioned, the systematic bias detected is extremely small. However, future larger GWAS with the statistical power to detect very small effects may need to control further the residual bias of certain confounders.

In conclusion, we provide the first comprehensive evidence of polygenicity patterns among human traits and diseases in EAS and EUR. Our findings confirm that there is an overall similarity between ancestry groups in the polygenicity of complex phenotypes. However, we observed ancestry-specific patterns in the polygenicity observed across multiple health domains. We hypothesize that these are due to evolutionary mechanisms that acted specifically in certain population groups. These may have important implications in the future applications of PGS to individuals of diverse ancestral backgrounds. Although these are novel insights into the genetics of complex traits, we have to acknowledge two main limitations. First, our study compared two ancestry groups leveraging data from three large biobanks. Because of the differences in the characteristics and the assessment of the cohorts investigated, our results may be partially due to cohort-specific factors rather than due to genetic variation between ancestries. Our study was based on publicly available datasets and we were not able to find additional resources to expand our analyses to other cohorts. Our findings will need to be confirmed in other large cohorts including diverse participants. Second, the difference in the sample size available for EAS and EUR may have affected the results of some of the analyses conducted. We hope that in the next years, additional data will become available for EAS and other population groups that are currently underrepresented in genetic research.

## MATERIALS and METHODS

### Study Populations

In our study, we used data generated from EAS and EUR participants enrolled in BBJ, UKB, and FinnGen. BBJ is a prospective biobank that collected DNA and serum samples from 12 medical institutions in Japan and recruited approximately 200,000 participants, mainly of Japanese ancestry. The mean age of participants at recruitment was 63 years old, and 46% were female. BBJ phenotype information including disease endpoints, past medical history, electronic health records (EHR), biomarkers, and prescription category (9). UKB is a biobank that enrolled more than 500,000 participants assessed through detailed web-based questionnaires on their diet, cognitive function, work history, health status, and other relevant phenotypes (10). EHRs are available for the UKB cohort, providing information regarding primary care, hospital episodes, death registry, and laboratory test results (10). The mean age of UKB participants at recruitment was 57 years old, and 54% were female. FinnGen is a public–private partnership project combining genotype data from Finnish biobanks and digital health record data from Finnish health registries (11). The mean age of participants at DNA sample collection was 52 years old and 56% were female.

The BBJ, UKB, and FinnGen genome-wide associations were generated following the same analytic approach described previously (12). Briefly, the association analysis of binary traits (i.e., disease endpoints and medication usage) was performed by using the generalized linear mixed model implemented SAIGE (v.0.37) (35), including age, age^2^, sex, age×sex, age^2^×sex and the top-20 principal components as covariates. For sex-specific diseases, age, age^2^, and the top-20 within ancestry principal components were included as covariates, and only controls of the sex to which the disease is specific were used. BOLT-LMM (v.2.3.4) (36) was used to conduct GWAS of quantitative traits (i.e., biomarkers) by using a linear mixed model and including the same covariates as used in the binary traits above. UKB and FinnGen GWAS meta-analyzed using the inverse-variance method to create a single EUR dataset (12). A total of 215 traits and 152 matching phenotypes were investigated in EAS and EUR, respectively (Supplemental Table 1).

### Effect size distribution

The GENESIS R package (5) was used to determine the descriptive statistics regarding the effect size distribution of complex traits in EAS and EUR (Supplementary Table 1). Briefly, GENESIS approach can distinguish susceptibility SNPs (i.e., those that have a detectable influence without requiring genome-wide significance) from null SNPs (i.e., those that have no detectable effect on a trait) to estimate parameters describing the polygenic architecture of a trait: i) π_c,_ the proportion of susceptibility SNPs per trait; ii) σ^2^, the variance of non-null SNPs; and iii) a, the residual effects not captured by the variance of effect-sizes (e.g., population stratification, underestimated effects of extremely small effect size SNPs, and/or genomic deflation) (5). As recommended by the developers (5), the GWAS data used in the present study were filtered to include only HapMap3 SNPs with an ancestry-specific minor allele frequency ≥0.05 (37). Using the *preprocessing()* function, SNPs were also removed if: i) their effective sample sizes were less than 0.67 times the 90th percentile of the per-SNP sample size distribution; ii) they were within the major histocompatibility region (excluded because of its complex LD structure; and iii) they had extremely large effect sizes (per-SNP effect z-scores>80). After these quality control steps, the *genesis()* function was used to implement the two-component model, which assumes that the distribution of effects for non-null SNPs follows a single normal distribution. The same preprocessing and parameter estimation was performed for EAS and EUR datasets. EAS and EUR LD score reference panels were used with respect to the corresponding ancestry group (available at https://github.com/yandorazhang/GENESISasian and https://github.com/yandorazhang/GENESIS, respectively).

The expected proportion of genetic variance explained by susceptibility SNPs reaching genome-wide significance considering projected sample sizes equal to 1,000,000 and 5,000,000 by applying the *projection()* function in GENESIS (5).

### Statistical analyses

We tested within- and between-population differences of the descriptive statistics related to the polygenic architecture of complex traits using non-parametric tests. This permitted us to avoid issues related to the distribution of the variables investigated and to the presence of possible outliers. The KW test was used to compare differences across multiple groups (e.g., phenotypic categories) in a single analysis. To follow up KW results, we used the Dunn test to perform post-hoc pairwise comparisons. To account for the number of pairwise comparisons, we applied FDR multiple testing correction. KW and Dunn tests were performed using the rstatix R package.

## Data Availability

All data produced in the present work are contained in the manuscript

## Acknowledgments

The authors thank Biobank Japan, UK Biobank, and FinnGen participants and investigators. The authors also acknowledge grants from the National Institutes of Health (R33 DA047527, R21 DC018098, and RF1 MH132337), and the One Mind.

## Abbreviations

BBJ: Biobank Japan
EAS: East Asian
EHR: Electronic Health Records
EUR: European
FDR: False Discovery Rate
GENESIS: Genetic effect-size distribution inference from summary-level data
GV%: proportion of genetic variance explained by susceptibility SNPs projected to reach genome-wide significance
GWAS: Genome-Wide Association Studies
KW: Kruskal-Wallis
LD: Linkage Disequilibrium
PGS: Polygenic Score
UKB: UK biobank

## REFERENCES

1 Abdellaoui, A., Yengo, L., Verweij, K.J.H. and Visscher, P.M. (2023) 15 years of GWAS discovery: Realizing the promise. Am J Hum Genet, 110, 179–194.

2 Loh, P.R., Bhatia, G., Gusev, A., Finucane, H.K., Bulik-Sullivan, B.K., Pollack, S.J., Schizophrenia Working Group of Psychiatric Genomics, C., de Candia, T.R., Lee, S.H., Wray, N.R. et al. (2015) Contrasting genetic architectures of schizophrenia and other complex diseases using fast variance-components analysis. Nat Genet, 47, 1385–1392.

3 Johnson, R., Burch, K.S., Hou, K., Paciuc, M., Pasaniuc, B. and Sankararaman, S. (2021) Estimation of regional polygenicity from GWAS provides insights into the genetic architecture of complex traits. PLoS Comput Biol, 17, e1009483.

4 Uricchio, L.H. (2020) Evolutionary perspectives on polygenic selection, missing heritability, and GWAS. Hum Genet, 139, 5–21.

5 Zhang, Y., Qi, G., Park, J.H. and Chatterjee, N. (2018) Estimation of complex effect-size distributions using summary-level statistics from genome-wide association studies across 32 complex traits. Nat Genet, 50, 1318–1326.

6 Zeng, J., de Vlaming, R., Wu, Y., Robinson, M.R., Lloyd-Jones, L.R., Yengo, L., Yap, C.X., Xue, A., Sidorenko, J., McRae, A.F. et al. (2018) Signatures of negative selection in the genetic architecture of human complex traits. Nat Genet, 50, 746–753.

7 O’Connor, L.J., Schoech, A.P., Hormozdiari, F., Gazal, S., Patterson, N. and Price, A.L. (2019) Extreme Polygenicity of Complex Traits Is Explained by Negative Selection. Am J Hum Genet, 105, 456–476.

8 Wendt, F.R., Pathak, G.A., Overstreet, C., Tylee, D.S., Gelernter, J., Atkinson, E.G. and Polimanti, R. (2021) Characterizing the effect of background selection on the polygenicity of brain-related traits. Genomics, 113, 111–119.

9 Ishigaki, K., Akiyama, M., Kanai, M., Takahashi, A., Kawakami, E., Sugishita, H., Sakaue, S., Matoba, N., Low, S.K., Okada, Y. et al. (2020) Large-scale genome-wide association study in a Japanese population identifies novel susceptibility loci across different diseases. Nat Genet, 52, 669–679.

10 Bycroft, C., Freeman, C., Petkova, D., Band, G., Elliott, L.T., Sharp, K., Motyer, A., Vukcevic, D., Delaneau, O., O’Connell, J. et al. (2018) The UK Biobank resource with deep phenotyping and genomic data. Nature, 562, 203–209.

11 Kurki, M.I., Karjalainen, J., Palta, P., Sipila, T.P., Kristiansson, K., Donner, K.M., Reeve, M.P., Laivuori, H., Aavikko, M., Kaunisto, M.A. et al. (2023) FinnGen provides genetic insights from a well-phenotyped isolated population. Nature, 613, 508–518.

12 Sakaue, S., Kanai, M., Tanigawa, Y., Karjalainen, J., Kurki, M., Koshiba, S., Narita, A., Konuma, T., Yamamoto, K., Akiyama, M. et al. (2021) A cross-population atlas of genetic associations for 220 human phenotypes. Nat Genet, 53, 1415–1424.

13 Vicks, W.S., Lo, J.C., Guo, L., Rana, J.S., Zhang, S., Ramalingam, N.D. and Gordon, N.P. (2022) Prevalence of prediabetes and diabetes vary by ethnicity among U.S. Asian adults at healthy weight, overweight, and obesity ranges: an electronic health record study. BMC Public Health, 22, 1954.

14 Ramachandran, A., Snehalatha, C., Shetty, A.S. and Nanditha, A. (2012) Trends in prevalence of diabetes in Asian countries. World J Diabetes, 3, 110–117.

15 Spracklen, C.N., Horikoshi, M., Kim, Y.J., Lin, K., Bragg, F., Moon, S., Suzuki, K., Tam, C.H.T., Tabara, Y., Kwak, S.H. et al. (2020) Identification of type 2 diabetes loci in 433,540 East Asian individuals. Nature, 582, 240–245.

16 Silva, F., Weisskopf, A., Castillo, C., Murphy, C., Kingwell-Banham, E., Qin, L. and Fuller, D.Q. (2018) A tale of two rice varieties: Modelling the prehistoric dispersals of japonica and proto-indica rices. The Holocene, 28, 1745–1758.

17 Jiang, L. and Liu, L. (2015) New evidence for the origins of sedentism and rice domestication in the Lower Yangzi River, China. Antiquity, 80, 355–361.

18 Landini, A., Yu, S., Gnecchi-Ruscone, G.A., Abondio, P., Ojeda-Granados, C., Sarno, S., De Fanti, S., Gentilini, D., Di Blasio, A.M., Jin, H. et al. (2021) Genomic adaptations to cereal-based diets contribute to mitigate metabolic risk in some human populations of East Asian ancestry. Evol Appl, 14, 297–313.

19 Malaria Genomic Epidemiology Network. (2019) Insights into malaria susceptibility using genome-wide data on 17,000 individuals from Africa, Asia and Oceania. Nat Commun, 10, 5732.

20 Zhang, X., Witt, K.E., Banuelos, M.M., Ko, A., Yuan, K., Xu, S., Nielsen, R. and Huerta-Sanchez, E. (2021) The history and evolution of the Denisovan-EPAS1 haplotype in Tibetans. Proc Natl Acad Sci U S A, 118.

21 Koller, D., Wendt, F.R., Pathak, G.A., De Lillo, A., De Angelis, F., Cabrera-Mendoza, B., Tucci, S. and Polimanti, R. (2022) Denisovan and Neanderthal archaic introgression differentially impacted the genetics of complex traits in modern populations. BMC Biol, 20, 249.

22 De Lillo, A., D’Antona, S., Pathak, G.A., Wendt, F.R., De Angelis, F., Fuciarelli, M. and Polimanti, R. (2021) Cross-ancestry genome-wide association studies identified heterogeneous loci associated with differences of allele frequency and regulome tagging between participants of European descent and other ancestry groups from the UK Biobank. Hum Mol Genet, 30, 1457–1467.

23 Trumble, B.C., Jaeggi, A.V. and Gurven, M. (2015) Evolving the neuroendocrine physiology of human and primate cooperation and collective action. Philos Trans R Soc Lond B Biol Sci, 370, 20150014.

24 Saitou, M., Resendez, S., Pradhan, A.J., Wu, F., Lie, N.C., Hall, N.J., Zhu, Q., Reinholdt, L., Satta, Y., Speidel, L. et al. (2021) Sex-specific phenotypic effects and evolutionary history of an ancient polymorphic deletion of the human growth hormone receptor. Sci Adv, 7, eabi4476.

25 Bergey, C.M., Lopez, M., Harrison, G.F., Patin, E., Cohen, J.A., Quintana-Murci, L., Barreiro, L.B. and Perry, G.H. (2018) Polygenic adaptation and convergent evolution on growth and cardiac genetic pathways in African and Asian rainforest hunter-gatherers. Proc Natl Acad Sci U S A, 115, E11256–E11263.

26 Li, J., Hong, X., Mesiano, S., Muglia, L.J., Wang, X., Snyder, M., Stevenson, D.K. and Shaw, G.M. (2018) Natural Selection Has Differentiated the Progesterone Receptor among Human Populations. Am J Hum Genet, 103, 45–57.

27 Schaschl, H. and Wallner, B. (2020) Population-specific, recent positive directional selection suggests adaptation of human male reproductive genes to different environmental conditions. BMC Evol Biol, 20, 27.

28 Zeberg, H., Kelso, J. and Paabo, S. (2020) The Neandertal Progesterone Receptor. Mol Biol Evol, 37, 2655–2660.

29 Chekalin, E., Rubanovich, A., Tatarinova, T.V., Kasianov, A., Bender, N., Chekalina, M., Staub, K., Koepke, N., Ruhli, F., Bruskin, S. et al. (2019) Changes in Biological Pathways During 6,000 Years of Civilization in Europe. Mol Biol Evol, 36, 127–140.

30 Lewis, C.M. and Vassos, E. (2020) Polygenic risk scores: from research tools to clinical instruments. Genome Med, 12, 44.

31 Khera, A.V., Chaffin, M., Aragam, K.G., Haas, M.E., Roselli, C., Choi, S.H., Natarajan, P., Lander, E.S., Lubitz, S.A., Ellinor, P.T. et al. (2018) Genome-wide polygenic scores for common diseases identify individuals with risk equivalent to monogenic mutations. Nat Genet, 50, 1219–1224.

32 Duncan, L., Shen, H., Gelaye, B., Meijsen, J., Ressler, K., Feldman, M., Peterson, R. and Domingue, B. (2019) Analysis of polygenic risk score usage and performance in diverse human populations. Nat Commun, 10, 3328.

33 Ruan, Y., Lin, Y.F., Feng, Y.A., Chen, C.Y., Lam, M., Guo, Z., Stanley Global Asia, I., He, L., Sawa, A., Martin, A.R. et al. (2022) Improving polygenic prediction in ancestrally diverse populations. Nat Genet, 54, 573–580.

34 Zhao, Z., Fritsche, L.G., Smith, J.A., Mukherjee, B. and Lee, S. (2022) The construction of cross-population polygenic risk scores using transfer learning. Am J Hum Genet, 109, 1998–2008.

35 Zhou, W., Nielsen, J.B., Fritsche, L.G., Dey, R., Gabrielsen, M.E., Wolford, B.N., LeFaive, J., VandeHaar, P., Gagliano, S.A., Gifford, A. et al. (2018) Efficiently controlling for case-control imbalance and sample relatedness in large-scale genetic association studies. Nat Genet, 50, 1335–1341.

36 Loh, P.R., Tucker, G., Bulik-Sullivan, B.K., Vilhjalmsson, B.J., Finucane, H.K., Salem, R.M., Chasman, D.I., Ridker, P.M., Neale, B.M., Berger, B. et al. (2015) Efficient Bayesian mixed-model analysis increases association power in large cohorts. Nat Genet, 47, 284–290.

37 International HapMap, C., Altshuler, D.M., Gibbs, R.A., Peltonen, L., Altshuler, D.M., Gibbs, R.A., Peltonen, L., Dermitzakis, E., Schaffner, S.F., Yu, F. et al. (2010) Integrating common and rare genetic variation in diverse human populations. Nature, 467, 52–58.

